# COVID-19 illness, SARS-CoV2 infection, and subsequent suicidal ideation in the French nationwide population-based EpiCov cohort : a propensity score analysis of more than 50,000 individuals

**DOI:** 10.1101/2022.08.02.22278311

**Authors:** Camille Davisse-Paturet, Massimiliano Orri, Stéphane Legleye, Aline-Marie Florence, Jean-Baptiste Hazo, Josiane Warszawski, Bruno Falissard, Marie-Claude Geoffroy, Maria Melchior, Alexandra Rouquette, the EPICOV study group

**Affiliations:** Paris-Saclay University, UVSQ, Inserm, Center for Epidemiology and Population Health, Paris, France; McGill University, Department of Psychiatry, Montreal, Québec, Canada; Ensai, Bruz, France; Sorbonne University, Inserm, Pierre Louis institute of Epidemiology and Public Health, Paris, France; French Ministry of Solidarity and Health, Drees, Paris, France; APHP, Paris-Saclay University, Department of Epidemiology and Public Health, Le Kremlin-Bicêtre, France; McGill University, Department of Educational and Counselling Psychology, Québec, Canada

## Abstract

Symptomatic COVID-19 appears to be associated with suicidal ideation but longitudinal evidence is still scarce. SARS-CoV2-induced neurological damages might underline this association, but findings are inconsistent. We therefore investigated the association between COVID-19 disease and subsequent suicidal ideation in the general population, using both self-reported symptoms and serology as well as inverse probability weighting to draw as near as possible to the direct association.

Using data from the nationwide French EpiCov cohort, COVID-19 disease was assessed through 1) COVID-19 illness (self-reported symptoms of sudden loss of taste/smell or fever alongside cough, shortness of breath or chest oppression, between February and November 2020), and 2) SARS-CoV2 infection (Spike protein ELISA test screening in dried-blood-spot samples). Suicidal ideation was self-reported between December 2020 and July 2021. Inverse probability weighting with propensity scores was used as an adjustment strategy, leading to balanced sociodemographic and health-related factors between the exposed and non-exposed groups of both COVID-19 disease measures. Then, modified Poisson regression models were used to investigate the association of COVID-19 illness and SARS-CoV2 infection with subsequent suicidal ideation.

Among 52,050 participants from the EpiCov cohort, 1.68% [1.54% - 1.82%] reported suicidal ideation in the first half of 2021, 9.57% [9.24% – 9.90%] had a SARS-CoV2 infection in 2020 and 13.23% [12.86% – 13.61%] reported COVID-19 symptoms in 2020. COVID-19 illness in 2020 was associated with higher risks of subsequent suicidal ideation in the first half of 2021 (Relative Risk_ipw_ [CI95%]= 1.43 [1.20 – 1.69]) while SARS-CoV2 infection in 2020 was not (RR_ipw_ = 0.88 [0.69 – 1.12]).

If COVID-19 illness was associated with subsequent suicidal ideation, the exact role of SARS-CoV2 infection with respect to suicide risk has yet to be clarified. Psychological support should be offered to persons recovering from symptomatic COVID-19 in order to minimize suicidal ideation risk. Moreover, if such psychological support is to be implemented, serology status alone does not seem a relevant criterion to define persons who suffered from COVID-19 to prioritize.

## Introduction

Since the beginning of the COVID-19 pandemic, mental health specialists have raised concerns regarding the possibility of an increase in the risk of suicidal behaviors among persons recovering from COVID-19.[1-3] Regarding potential biological pathways, the exact role of SARS-CoV2 with regard to mechanisms involved in suicide risk has yet to be demonstrated. Nonetheless, the virus’s ability to invade the central nervous system through fixation on angiotensin converting enzyme 2 receptors,[4] or to inflict brain damage through hyperinflammation,[5] are potential candidates. From a public health point of view, suicide risk related to COVID-19 disease was first supported by findings from cross-sectional studies and case series,[6, 7] and limited evidence from longitudinal studies is now emerging. A systematic review ascertaining the risk of suicidal and self-harm thoughts and behaviors among persons recovering from SARS-CoV2 infection identified 11 relevant studies conducted between January 2020 and July 2021 and representing eight separate samples.[8] Eight out of the 11 studies reported elevated risk of suicidal or self-harm thoughts after SARS-CoV2 infection. Unfortunately, these studies were quite heterogenous in terms of suicide risk assessment, study population, or design. Of note, one had a longitudinal cohort design.[9] In this study, self-reported suspicion or diagnosis of COVID-19 was associated with an elevated risk of self-harm thoughts or behaviors over 59 weeks since then end of March 2020. A recent longitudinal study from Australia also reported an increased risk of suicidal ideation in the three months following exposure to COVID-19, even when controlling for the pandemic’s impact on employment and financial distress.[10] Yet, as explained by the authors, the situation in Australia at the time of assessment, between March and June 2020, was quite different from that in other parts of the world, including Europe. In a paper gathering data from seven longitudinal cohorts from six Northern-Europe countries, the number of days spent in bed due to SARS-CoV2 infection, was cross-sectionally associated to mental health outcomes in a dose-effect way.[11] Participants not bedridden by the infection were at lower risk of both depressive and anxiety symptoms than those who were not infected, while those bedridden for seven days or more were at higher risk. As depressive and anxiety symptoms are known trigger of suicidal behaviors,[12] taking into account both the infection status and its associated symptoms might therefore be relevant when studying suicide risk. Moreover, as both COVID-19 disease and suicidal behaviors are related to individuals’ sociodemographic and health characteristics,[12, 13] comprehensive information regarding theses aspects is needed to address the association between COVID-19 disease and suicidal behavior. To maximize such assessment and get one step closer to causal inference, the use of propensity scores seems promising as it ensures a balance of all selected covariates between exposure groups.[14] Our aim was therefore to study the long-term association between COVID-19 illness, as assessed by self-reported symptoms of COVID-19, SARS-CoV2 infection, as assessed by serology status, in 2020, and subsequent suicidal ideation in 2021. We used data from a French nationwide cohort and accounted for a wide range of sociodemographic and health-related factors through inverse probability weighting.

## Methods

### Study population

The Epidémiologie et conditions de vie sous le COVID-19 (EpiCov) study is a longitudinal, nationwide, French cohort aiming to provide information on the virus’ dissemination and the pandemic’s consequences on the daily life and health of individuals.[15] Eligibility criteria were to be at least 15 years of age in 2020, to reside in Metropolitan France or three oversea territories (Martinique, Guadeloupe and Réunion), and to not live in a medical retirement home or a jail. 371,000 individuals were randomly selected from France’s national tax database, with an expected participation rate of about 50% and a sampling design overrepresenting less densely populated and more socioeconomically disadvantaged areas.[16]

EpiCov participants were followed through three waves of data collection, using self-computer-assisted-web interviews (CAWI) or computer-assisted-telephone interviews (CATI). From the 371,000 randomly selected individuals, 36.22% (134,391) participated in the first wave (02/05/2020 – 02/06/2020, later referred to as baseline). Of these 134,391 individuals, 80.18% (107,759) participated the second wave of data collection (26/10/2020 – 14/12/2020), and 63.30% (85,074) were still followed in the third wave of data collection (24/06/2021 – 09/08/2021). The EpiCov study timeline as well as data collected and used at each follow-up wave are available in supplementary figure 1.

**Figure 1:**
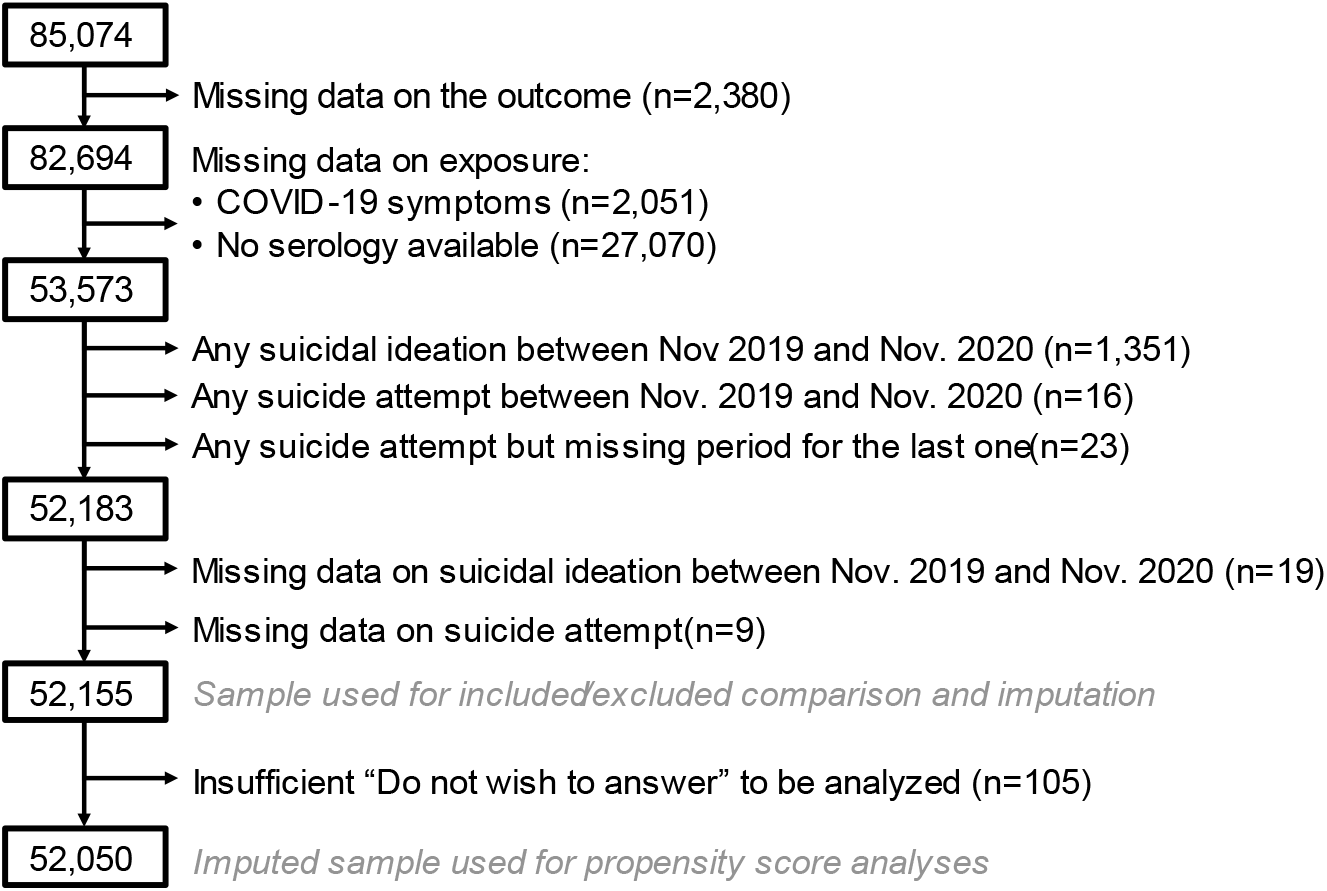
study sample selection from the 85,074 individuals who participated to the three follow-up waves of the EpiCov cohort, in 2020 and 2021, in France.

The EpiCov study received approval from an ethics committee (Comité de Protection des Personnes Sud Méditerranée III 2020-A01191-38) and from France’s National Data Protection Agency (Commission Nationale Informatique et Libertés, CNIL, MLD/MFI/AR205138).

### Outcome: suicidal ideation between December 2020 and July 2021

Occurrence of suicidal ideation from December 2020 to July 2021 (yes vs no) was ascertained during the third follow-up wave as follows: “since December 2020, have you thought about committing suicide?” (yes vs no).

### Exposures

#### COVID-19 infection: SARS-CoV2 serology

The exact methodology for serology testing has been described elsewhere.[15] Briefly, among 85,350 individuals who participated in the second follow-up wave and who agreed to receive a blood sampling kit, 66,826 dried-blood-spot samples were returned and 63,524 were screened for antibodies against SARS-CoV2’s spike protein S1 domain, with the use of a commercial ELISA kit. Samples with an optical density ratio above 1.1 were considered as positive for SARS-CoV2 testing, while a ratio between 0.7 and 1.1 was considered as suspicious. As a decline in circulating antibodies might occur with time[17], both positive and suspicious serologies were considered as markers of SARS-CoV2 infection in this study. Positive serological status was unlikely to be due to vaccination as more than 90% of the dried-blood-spot samples used in the present study were performed before January 2021 and the start of the vaccination campaign in France.

#### COVID-19 illness: self-reported symptoms between February 2020 and November 2020

As a wide range of symptoms can be associated with a SARS-CoV2 infection, only symptoms described as most suspicious in 2020 by the French Public Health Agency were ascertained. Using data from the baseline and the second follow-up wave, the occurrence of COVID-19 symptoms (yes or no) between February and November 2020 was defined as a self-report of any unusual episode of sudden loss of taste/smell or any unusual episode of fever alongside a cough, shortness of breath, or chest oppression.

### Covariates for propensity score model

Covariate selection for propensity score modeling was based on current literature and knowledge about factors involved in both COVID-19 and suicidal ideation risks. Of note, particular attention was given to covariates related to suicidal ideation.[7] Directed acyclic graphs (DAGs) were implemented to help conceptualize a framework for the assessment of suicidal ideation related to COVID-19 and minimize bias though appropriate covariate selection.[18, 19]

### Sociodemographic and health covariates

The following sociodemographic covariates were ascertained at baseline and included in propensity scores: gender (man, woman), age (years), participant’s and participant’s parents’ place of birth (both participant and parents born in mainland France, participant or parents born in oversea territories, participant born in France of parents born abroad, participant born abroad), highest educational attainment (none, lower secondary school certificate, professional certificate, higher secondary school certificate, bachelor degree or equivalent, Master degree or more), occupational grade (employed, student, unemployed, retired, other including housemakers), perceived financial situation (comfortable, decent, short, difficult or unbearable without taking loans), less than one room per person in participant’s usual accommodation (yes or no), residence not in usual housing during the first lockdown (yes or no), access to a private exterior during the first lockdown (balcony or garden, including common ones, yes or no), and usual living area according to the intensity of the first COVID-19 wave in France (less affected areas, Grand-Est, Hauts-de-France, Ile-de-France).

The following health-related covariates were also ascertained: perceived general health status at baseline (very good to good, quite good, poor to very poor), baseline body mass index (BMI, less than 18.5 kg/m^2^, between 18.5 and less than 25 kg/m^2^, between 25 and less than 30 kg/m^2^, 30kg/m^2^ or more), pre pandemic somatic conditions (yes or no), pre pandemic mental health disorder (yes or no), baseline tobacco use (current, past, never) and baseline alcohol use (daily, often, occasional, rare, never). As pre pandemic mental health disorder is a key factor when studying suicidal ideation, data from the second and third follow-up waves were used to complete baseline information. This covariate hence includes self-reported anxiety, depression and mental disability, or history of at least one suicide attempt before November 2019, or self-report of at least one physician diagnosis of anxiety, mood, bipolar, eating, personality or substance use disorder or schizophrenia before the pandemic.

### Available indicators

The following indicators, made available by The National Institute for Statistics and Economic Studies (INSEE), were taken into account: deciles of household income per consumption unit studied as a five-category covariate (less resourceful, medium-low, medium, medium-high, wealthiest), household structure (single, couple without children, couple with children, single-parent, participant living with parents, complex household), urban density of living area (oversea territories, less than 2,000 urban units, between 2,000 and 1,999,999 urban units, Paris area), residence in a deprived neighborhood (yes or no), and hospitalization rates in place of residence during the first lockdown (lowest, medium-low, medium-high, highest quartile). An urban unit is a built area with less than 200 meters between two buildings, comprising at least 2,000 inhabitants. A deprived neighborhood is an administrative category to identify an area where particular budgetary efforts are made by the State to tackle inequalities regarding education, early life care, housing and living conditions, employment, social cohesion, security, and crime prevention.

### Statistical analyses

Suicidal ideation in the past 12 months, as well as life course suicide attempts were assessed in the second follow-up wave (supplementary figure 1). The related questions were respectively: “In the last 12 months, have you thought about committing suicide?” and “In your lifetime, have you ever attempted suicide?”. To ensure COVID-19 illness or SARS-CoV2 infection actually happened before suicidal ideation, participants reporting suicidal ideation in the last 12 months or a history of suicide attempt between November 2019 and 2020, or who did not provide information on the timing of their last suicide attempt, were excluded from the statistical analyses. The number of participants excluded at each step of selection are detailed in Figure 1.

### Descriptive statistics

Study weights were applied to all descriptive statistics in order to take EpiCov’s design and attrition bias into account. Briefly, the study weights accounted for demographic and socioeconomic indicators potentially linked to response probability and made available by INSEE from the tax data base. Study weights were also calibrated on margins of the general population according to census data and population projections.[20]

### Inverse probability weighting and modified Poisson regression models

First, propensity scores associated with a) reporting COVID-19 symptoms or b) having a positive SARS-CoV2 serology were computed using logistic regression models based on the covariates described above. Then, propensity scores were included in statistical models using inverse probability weights (IPWs). Balance after IPWeighting was considered satisfactory if: 1) absolute standardized mean differences (SMDs) between each covariate, as well as each category of each covariate were below 10%, 2) variance ratios of propensity scores computed after weighting were between 0.5 and 2.[14, 21] Covariates distribution after weighting was also assessed with chi-squared and student t-tests. Lastly, IPWeighted modified Poisson regression models with robust error variance were used to assess the association between COVID-19 infection and suicidal ideation. [22] If after IPWeighting residual distribution differences remained for some covariates, regression models were further adjusted for these incompletely balanced covariates. Models were therefore further adjusted for highest educational attainment when assessing COVID-19 illness, and for highest educational attainment, perceived financial situation, household income, and residence in deprived neighborhood when assessing SARS-CoV2 infection.

### Sensitivity analyses

To test the robustness of our results, we also estimated relative risks (RR) by weighting the reference groups (no COVID-19 illness; no SARS-CoV2 infection) to match the covariates’ distribution in the exposed groups (respectively: COVID-19 illness; SARS-CoV2 infection). This method, called average treatment effect on the treated (ATT), assesses what would have happened to COVID-19 ill participants, or seropositive to SARS-CoV2 participants, if they had not been ill or infected. The more consistent the estimated RRs in IPWeighted and ATTWeighted analyses, the more robust the results.

Although we accounted for a wide range of covariates in propensity scores calculation, the probability of not having a comprehensive list of all relevant confounding factors cannot be ruled out. To control for this probability we estimated E-values.[23] An E-value gives the value of the joint minimum strength of association an unmeasured confounder must have with both the exposure and outcome to fully explain the association found between the exposure and outcome, after adjusting for the measured covariates. In the present study, the E-value is given on the relative risk scale. The higher the E-value is, the more strength a potential unmeasured confounder needs to explain the association between the exposure and outcome and the less likely it is to exist.

Interactions between gender, or age, and the two COVID-19 exposures with respect to subsequent suicidal ideation were assessed but did not reached statistical significance (all p-value above 0.20).

All the analyses were performed using SAS V9.4. Tests were two-sided and considered statistically significant at p < 0.05. Missing data on covariates (up to 4.84%) were handled using the fully conditional specification method and assuming that data were missing at random (SAS MI procedure, FCS statement). After imputation, one imputed data set was randomly selected for propensity score analyses.

## Results

### Description of the study sample

The final sample included 52,050 participants. As reported in Table 1, our study sample was mostly composed of female, over 25 years of age, born in mainland France, living with a partner with or without children, employed or retired, with at least a professional certificate, living in wealthiest households, and with decent to short perceived financial situations. They were less likely to live in oversea territories or the Paris area, in accommodations with less than one room per person, and in a deprived neighborhood. The study sample participants also more often reported feeling good to very good, had no chronic condition, were non-smokers and non or occasional alcohol drinkers. Most of them resided in their usual accommodation during the first lockdown (17/03/2020 – 11/05/2020), had access to a private exterior, and were more likely to live in an area less affected by the first COVID-19 wave.

**Table 1:** outcome, exposures, and covariates distribution in study sample, weighted by study weights, n = 52,050

|  | Distribution |
| --- | --- |
| *Suicidal ideations since December 2020* |  |
| No | 98.32% [98.18 - 98.46] |
| Yes | 1.68% [1.54 - 1.82] |
| *SARS-CoV2 infection in 2020* |  |
| No | 90.43% [90.1 - 90.76] |
| Yes | 9.57% [9.24 - 9.90] |
| *Covid-19 illness between February and December 2020* |  |
| No | 86.77% [86.39 - 87.14] |
| Yes | 13.23% [12.86 - 13.61] |
| *Gender* |  |
| Men | 46.16% [45.58 - 46.73] |
| Women | 53.84% [53.27 - 54.42] |
| *Age (years)* |  |
| 15 - 25 | 11.25% [10.90 - 11.61] |
| 26 - 45 | 27.83% [27.33 - 28.33] |
| 46 - 65 | 34.92% [34.40 - 35.44] |
| > 65 | 26.00% [25.44 - 26.56] |
| *Place of birth* |  |
| Both participant and parents born in mainland France | 82.01% [81.53 - 82.50] |
| Participant or parents born in oversea territories | 1.96% [1.81 - 2.11] |
| Participant born in France of parents born abroad | 8.61% [8.27 - 8.95] |
| Participant born abroad | 7.41% [7.05 - 7.78] |
| *Highest educational attainment* |  |
| None | 8.20% [7.75 - 8.65] |
| Lower secondary school certificate | 13.31% [12.83 - 13.78] |
| Professional certificate | 19.11% [18.66 - 19.56] |
| Baccalauréat (higher secondary school certificate) | 19.55% [19.12 - 19.98] |
| Bachelor degree or equivalent | 25.34% [24.90 - 25.78] |
| Master degree or more | 14.49% [14.16 - 14.83] |
| *weighted % [95% Confidence interval]* |  |

Table 1 continued :
|  | Distribution |
| --- | --- |
| *Occupational grade* |  |
| Employed | 48.02% [47.45 - 48.59] |
| Students | 8.75% [8.45 - 9.06] |
| Unemployed | 4.63% [4.36 - 4.90] |
| Retired | 30.36% [29.79 - 30.92] |
| Other including housemakers | 8.23% [7.89 - 8.58] |
| *Perceived financial situation* |  |
| Comfortable | 16.15% [15.78 - 16.52] |
| Decent | 43.37% [42.80 - 43.93] |
| Short | 31.02% [30.46 - 31.58] |
| Difficult or unbearable without taking loans | 9.47% [9.07 - 9.87] |
| *Household structure* |  |
| Single | 16.73% [16.23 - 17.22] |
| Couple without children | 32.12% [31.59 - 32.65] |
| Couple with children | 28.75% [28.26 - 29.23] |
| Single-parent | 7.21% [6.90 - 7.52] |
| Participant living with parents | 7.85% [7.55 - 8.16] |
| Complex household | 7.34% [7.04 - 7.65] |
| *Household income per consumption units* |  |
| Less resourceful | 13.79% [13.33 - 14.25] |
| Medium-low | 16.60% [16.09 - 17.11] |
| Medium | 19.85% [19.38 - 20.32] |
| Medium-high | 23.82% [23.37 - 24.28] |
| Wealthiest | 25.93% [25.50 - 26.36] |
| *Less than one room per person in usual accommodation* |  |
| No | 93.32% [93.00 - 93.64] |
| Yes | 6.68% [6.36 - 7.00] |
| *Residence not in usual housing during the first lockdown* |  |
| No | 95.03% [94.78 - 95.27] |
| Yes | 4.97% [4.73 - 5.22] |
| *Access to a private exterior during the first lockdown* |  |
| No | 8.67% [8.31 - 9.03] |
| Yes | 90.40% [90.03 - 90.78] |
| Other situations | 0.93% [0.80 - 1.05] |
| *Usual living area* |  |
| Less affected area | 65.67% [65.13 - 66.21] |
| Grand-Est | 8.70% [8.39 - 9.01] |
| Hauts-de-France | 8.34% [8.02 - 8.67] |
| Ile-de-France | 17.29% [16.86 - 17.72] |
| *Urban density of living area (urban units)* |  |
| Oversea territories | 1.26% [1.15 - 1.37] |
| Less than 2,000 | 24.07% [23.58 - 24.55] |
| Between 2,000 and 1,999,999 | 59.58% [59.02 - 60.14] |
| Paris area | 15.09% [14.68 - 15.50] |
| *Usual residence in deprived neighborhood* |  |
| No | 95.69% [95.39 - 95.99] |
| Yes | 4.31% [4.01 - 4.61] |
| *weighted % [95% Confidence interval]* |  |

Table 1 continued :
|  | Distribution |
| --- | --- |
| *Perceived general health status* |  |
| Good to very good | 77.80% [77.27 - 78.34] |
| Quite good | 18.70% [18.21 - 19.20] |
| Poor to very poor | 3.49% [3.23 - 3.75] |
| *Body Mass Index (kg/m<sup>2</sup>)* |  |
| Under 18.5 | 3.38% [3.18 - 3.58] |
| From 18.5 to under 25 | 50.80% [50.23 - 51.38] |
| From 25 to under 30 | 30.89% [30.36 - 31.43] |
| From 30 and over | 14.93% [14.49 - 15.36] |
| *Pre pandemic somatic condition* |  |
| None | 60.91% [60.33 - 61.48] |
| At least one | 39.09% [38.52 - 39.67] |
| *Pre pandemic mental health disorder* |  |
| None | 89.00% [88.62 - 89.38] |
| At least one | 11.00% [10.62 - 11.38] |
| *Tobacco use* |  |
| Never | 48.76% [48.18 - 49.33] |
| Past only | 32.32% [31.79 - 32.84] |
| Current | 18.93% [18.46 - 19.39] |
| *Alcohol use* |  |
| Never | 29.39% [28.83 - 29.94] |
| Rare | 13.59% [13.19 - 13.99] |
| Occasional | 23.35% [22.88 - 23.82] |
| Often | 23.12% [22.67 - 23.57] |
| Daily | 10.56% [10.21 - 10.91] |
| *Hospitalization rates in place of residence during the 1st lockdown* |  |
| Lowest | 23.63% [23.14 - 24.12] |
| Medium-low | 27.89% [27.38 - 28.41] |
| Medium-High | 22.56% [22.08 - 23.04] |
| Highest | 25.92% [25.42 - 26.41] |
| *weighted % [95% Confidence interval]* |  |

### COVID-19 disease and subsequent suicidal ideation

Amongst the 52,050 participants with no history of suicidal ideation nor suicide attempts in 2020, 1.68% [1.54% – 1.82%] (863) reported suicidal ideation between December 2020 and July 2021, 9.57% [9.24% – 9.90%] (5,098) had a serology-confirmed SARS-CoV2 infection and 13.23% [12.86% – 13.61%] (7,058) reported COVID-19 symptoms between February and November 2020 (Table 1). Using unadjusted modified Poisson regression model, COVID-19 symptoms were associated with a higher risk of suicidal ideation (RR [CI95%]: 1.90 [1.63 – 2.23]) while the same association was not observed for serology-confirmed SARS-CoV2 infection (0.98 [0.78 – 1.23]).

As shown in Figure 2, these results remained unchanged after taking covariates into account with IPWeighting. Participants reporting COVID-19 symptoms had almost a 1.5 fold increased risk of subsequent suicidal ideation (RR_ipw_: 1.43 [1.20 – 1.69]) while those with a positive serology were not at risk (RR_ipw_: 0.88 [0.69 – 1.12]). Sensitivity ATTWeighted analyses yielded similar results (RR_watt_: 1.43 [1.22 – 1.68] for COVID-19 symptoms; RR_watt_: 0.94 [0.75 – 1.18] for serology-confirmed SARS-CoV2 infection).

**Figure 2:**
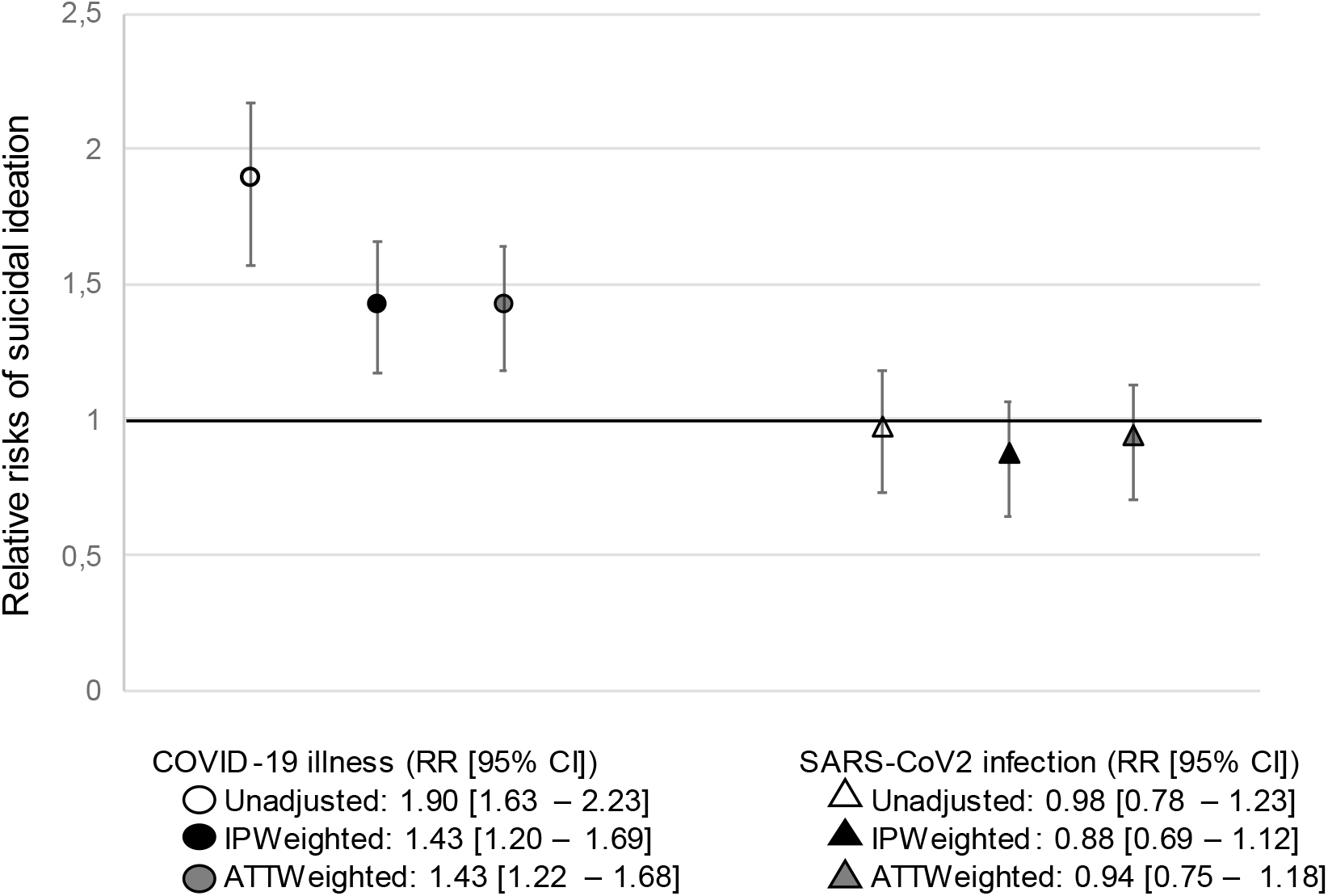
relative risks of suicidal ideation in 2021 in COVID-19 ill or SARS-CoV2 infected participants in 2020 from the EpiCov cohort, France

The minimum strength of association a potential unmeasured confounder should have with both COVID-19 illness and subsequent suicidal ideation after IPWeighting is given by the E-value: 2.21 [1.69 - 2.78]. It means that the observed RR_ipw_ of 1.43 would be completely explained by an unmeasured confounder associated with both COVID-19 illness and subsequent suicidal ideation with a relative risk of 2.21, after controlling for the measured covariates. However, given the number of measured confounders taken into account, such a strong unobserved confounder is rather unlikely.

## Discussion

### Summary of findings

In a nationwide study from France, COVID-19 illness in 2020, as defined by self-reports of sudden loss of taste/smell or fever alongside cough or shortness of breath or chest oppression, was associated with an almost 1.5 higher risk of subsequent suicidal ideation in the first half of 2021. Associations persisted after adjusting for a wide range of sociodemographic and health-related factors, using inverse probability weighting. However, SARS-CoV2 status, as confirmed by circulating antibodies, was not associated with subsequent suicidal ideation. To the best of our knowledge it is the first time the causal association between COVID-19 illness, SARS-CoV2 infection and subsequent suicidal ideation is assessed in such a large, randomly selected, longitudinal study.

### SARS-CoV2 infection and suicidal ideation

Our findings suggest that SARS-CoV2 is not likely to be involved in suicidal ideation. Although misclassification of infected individuals with very low levels of circulating antibodies cannot be ruled out, misclassification due to a new variant impairing effective detection of antibodies in blood samples seems unlikely as the first known variant of concern, the alpha variant, was first observed in France at the very end of 2020.[24] In line with our finding, the meta-analysis of 5 European cohorts highlighted that individuals with a COVID-19 diagnosis but less acute COVID-19 symptoms were at lower risk of depressive and anxiety symptoms than individuals without a COVID-19 diagnosis.[11] To dive further into the role of SARS-CoV2 in the etiology of suicide risk, future studies should maybe assess suicide risk with respect to the biological consequences of the infection, such as the presence of inflammation processes, rather than only the presence of the virus.[5]

### COVID-19 illness and suicidal ideation

Several mechanisms can explain the association between COVID-19 reported symptoms and suicidal ideation. First, some participants with COVID-19 required hospitalization, sometimes in traumatic units such as intense care ones,[25] and some had persistent physical symptoms.[26] A possible mechanism could be that such stressful and exhausting situations can have negative impacts on the quality of life of individuals, leading to higher risk of suicidal ideation. As an example, long-lasting symptoms or hospitalization could have impaired employment which may lead to financial distress, a risk factor of suicidal ideation.[9, 10] Second, anxiety and depressive symptoms could act as mediators of the relationship between COVID-19 and suicidal ideation. Indeed, COVID-19 has been found to be associated with a higher risk of depression,[9, 11, 25] a predictor of suicidal behaviors.[12] Third, Paul and Fancourt showed that COVID-19 illness or death among friends/family or closed ones was associated with a higher risk of self-harm thoughts and behaviors.[9] This was also true for worries about relatives in the preceding week. As COVID-19 is a communicable disease, individuals with symptomatic COVID-19 could be more likely to have symptomatic cases among their relatives, increasing their risk of suicidal ideation, especially if they feel responsible for their relatives’ infection.

### Strengths and limitations

A main limitation of our study is the lack of prospectively collected pre-pandemic information as the EpiCov study was initiated in 2020. Nonetheless, many pre-pandemic characteristics were collected retrospectively and could be taken into account. Specific attention was given to reports of previous mental health disorders, as they are key predictors of suicidal behaviors.[12] We considered participants’ history of anxiety, depression, or mental impairment, history of a suicide attempt before November 2019, and self-report of a physician diagnosis of psychiatric disorders. Although less accurate than prospectively collected or health-record data, these information give valuable insights of pre-pandemic mental health conditions.

Regarding propensity scores, balance between persons who did or did not experience COVID-19 is only achieved for covariates included in propensity score estimations. Definition of relevant factors to assess COVID-19 disease and suicidal ideation was based on the existing scientific literature and availability of information in study questionnaires. The probability of imperfect balance due to unmeasured factors cannot be ruled out. Yet, many factors were taken into account, quality of propensity score weighting was systematically assessed and available as supplementary material, and the estimated E-value of 2.21 [1.69 - 2.78] indicated a low probability of an unmeasured factor completely explaining the observed association between COVID-19 illness and subsequent suicidal ideation.

## Conclusion

Self-reported COVID-19 illness but not serology-confirmed SARS-CoV2 infection was associated with a higher risk of subsequent suicidal ideation while adjusting for a wide range of sociodemographic and health-related factors using inverse probability weighting. To highlight relevant targets for intervention, future studies should explore the long-term impact of symptomatic COVID-19, both biologically confirmed and not confirmed, on the quality of life of individuals as serology status alone do not seem like a relevant one.

## Data Availability

Anonymous aggregated data for the baseline wave are available online. The EpiCov dataset is available for research purpose concerning baseline wave, and will be available by March 2022 concerning the second follow-up wave for research purpose on CASD (https://www.casd.eu/), after submission to approval of French Ethics and Regulatory Committee procedure (Comite du Secret Statistique, CESREES and CNIL). Access to anonymized individual data underlying the findings may be available before the planned period, on request to the corresponding author, to be submitted to approval of ethics and reglementary Committee for researchers who meet the criteria for access to data.

## Authors contribution

CDP: conceptualisation, data curation, formal analysis, methodology, project administration, validation, visualisation, writing - original draft

MO: conceptualisation, funding acquisition, supervision, validation, writing - review & editing

SL: methodology, supervision, validation, writing - review & editing

AMF: conceptualisation, data curation, validation, writing - review & editing

JBH: conceptualisation, data curation, investigation, validation, writing - review & editing

JW: investigation, validation, writing - review & editing

BF: methodology, supervision, validation, writing - review & editing

MCG: conceptualisation, funding acquisition, supervision, validation, writing - review & editing

MM: conceptualisation, methodology, supervision, validation, writing - review & editing

AR: conceptualisation, funding acquisition, methodology, project administration, supervision, validation, writing - review & editing

## Data sharing

Anonymous aggregated data for the baseline wave are available online. The EpiCov dataset is available for research purpose concerning baseline wave, and will be available by March 2022 concerning the second follow-up wave for research purpose on CASD (https://www.casd.eu/), after submission to approval of French Ethics and Regulatory Committee procedure (Comité du Secret Statistique, CESREES and CNIL). Access to anonymized individual data underlying the findings may be available before the planned period, on request to the corresponding author, to be submitted to approval of ethics and reglementary Committee for researchers who meet the criteria for access to data.

## Acknowledgment

The present work was supported by a French National Observatory of Suicide research grant attributed to AR (grant number R21094LL). The EpiCov study was supported by research grants from Inserm (Institut National de la Santé et de la Recherche Médicale), the French Ministry for Research and its department of research, studies, evaluation and statistics (Direction de la Recherche, des Etudes, de l’Evaluation et des Statistiques, Drees), the French Ministry for Health, and the Région Ile de France. Dr.Bajos has received funding from the European esearch Council E C) under the European nion’s Hori on 2020 research and innovation program (grant agreement number 856478). This project has also received funding from the European nion’s Hori on 2020 research and innovation program under grant agreement number 101016167, ORCHESTRA (Connecting European Cohorts to Increase Common and Effective Response to SARS-CoV-2 Pandemic).

For the purpose of Open Access, a CC-BY public copyright licence has been applied by the authors to the present document and will be applied to all subsequent versions up to the Author Accepted Manuscript arising from this submission.

[CC-BY 4.0](https://creativecommons.org/licenses/by/4.0/)

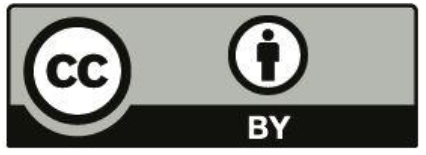

## Reference

1. Gunnell, D., et al., Suicide risk and prevention during the COVID-19 pandemic. Lancet Psychiatry, 2020. 7(6): p. 468–471.

2. Conejero, I., et al., [Suicidal behavior in light of COVID-19 outbreak: Clinical challenges and treatment perspectives]. Encephale, 2020. 46(3s): p. S66–s72.

3. Sher, L., Post-COVID syndrome and suicide risk. Qjm, 2021. 114(2): p. 95–98.

4. Szcześniak, D., et al., The SARS-CoV-2 and mental health: From biological mechanisms to social consequences. Prog Neuropsychopharmacol Biol Psychiatry, 2021. 104: p. 110046.

5. Costanza, A., et al., Hyper/neuroinflammation in COVID-19 and suicide etiopathogenesis: Hypothesis for a nefarious collision? Neurosci Biobehav Rev, 2022. 136: p. 104606.

6. John, A., et al., The impact of the COVID-19 pandemic on self-harm and suicidal behaviour: update of living systematic review. F1000Res, 2020. 9: p. 1097.

7. Hazo, J.-B., et al., Une dégradation de la santé mentale chez les jeunes en 2020 - Résultats issus de la 2e vague de l’enquête EpiCov. Etudes & Résultats, 2021. 1210.

8. Sinyor, M., et al., SARS-CoV-2 Infection and the Risk of Suicidal and Self-Harm Thoughts and Behaviour: A Systematic Review. Can J Psychiatry, 2022: p. 7067437221094552.

9. Paul, E. and D. Fancourt, Factors influencing self-harm thoughts and behaviours over the first year of the COVID-19 pandemic in the UK: longitudinal analysis of 49 324 adults. Br J Psychiatry, 2022. 220(1): p. 31–37.

10. Batterham, P.J., et al., Effects of the COVID-19 pandemic on suicidal ideation in a representative Australian population sample-Longitudinal cohort study. J Affect Disord, 2022. 300: p. 385–391.

11. Magnúsdóttir, I., et al., Acute COVID-19 severity and mental health morbidity trajectories in patient populations of six nations: an observational study. Lancet Public Health, 2022. 7(5): p. e406–e416.

12. Turecki, G. and D.A. Brent, Suicide and suicidal behaviour. Lancet, 2016. 387(10024): p. 1227–39.

13. Allen, J., et al., COVID-19 and the social determinants of health and health equity. 2021, World Health Organization: Geneva. p. 32.

14. Austin, P.C. and E.A. Stuart, Moving towards best practice when using inverse probability of treatment weighting (IPTW) using the propensity score to estimate causal treatment effects in observational studies. Stat Med, 2015. 34(28): p. 3661–79.

15. Warszawski, J., et al., Prevalence of SARS-Cov-2 antibodies and living conditions: the French national random population-based EPICOV cohort. BMC Infect Dis, 2022. 22(1): p. 41.

16. Statistique, C.N.d.l.I. EpiCoV : Étude EPIdémiologique de la diffusion du SARS-CoV2 - Vague T1 - 2020X711SA. 2020 [cited 2022 May 2022]; Available from: https://www.cnis.fr/enquetes/epicov-etude-epidemiologique-de-la-diffusion-du-sars-cov2-2020x711sa/.

17. Warszawski, J., et al., 4% de la population a développé des anticorps contre le SARS-CoV-2 entre mai et novembre 2020, in Etudes et Résultats, d.E. Direction de la Recherche, de l’Evalutation et des Statistiques, Editor. 2021, French Ministry of Health: France. p. 8.

18. Shrier, I. and R.W. Platt, Reducing bias through directed acyclic graphs. BMC Med Res Methodol, 2008. 8: p. 70.

19. Hernan, M. and J.M. Robins, Causal Inference: What if? 2020.

20. Warszawski, J., et al., A national mixed-mode seroprevalence random population-based cohort on SARS-CoV-2 epidemic in France: the socio-epidemiological EpiCov study. medRxiv, 2021: p. 2021.02.24.21252316.

21. Rubin, D.B., The design versus the analysis of observational studies for causal effects: parallels with the design of randomized trials. Stat Med, 2007. 26(1): p. 20–36.

22. Zou, G., A modified poisson regression approach to prospective studies with binary data. Am J Epidemiol, 2004. 159(7): p. 702–6.

23. VanderWeele, T.J. and P. Ding, Sensitivity Analysis in Observational Research: Introducing the E-Value. Ann Intern Med, 2017. 167(4): p. 268–274.

24. France, S.P. Coronavirus : circulation des variants du SARS-CoV-2. 2022 11/05/2022 [cited 2022 16/05/2022]; Available from: https://www.santepubliquefrance.fr/dossiers/coronavirus-covid-19/coronavirus-circulation-des-variants-du-sars-cov-2.

25. Taquet, M., et al., 6-month neurological and psychiatric outcomes in 236 379 survivors of COVID-19: a retrospective cohort study using electronic health records. Lancet Psychiatry, 2021. 8(5): p. 416–427.

26. Sykes, D.L., et al., Post-COVID-19 Symptom Burden: What is Long-COVID and How Should We Manage It? Lung, 2021. 199(2): p. 113–119.

