## Supplemental Material for "COVID-19 illness, SARS-CoV2 infection, and subsequent suicidal ideation in the French nationwide population-based EpiCov cohort : a propensity score analysis of more than 50,000 individuals"

***<u>Supplementary Material</u>***

### *The EpiCov study timeline*

*Supplementary Figure 1 : the EpiCov study timeline and data used at each follow -up wave*

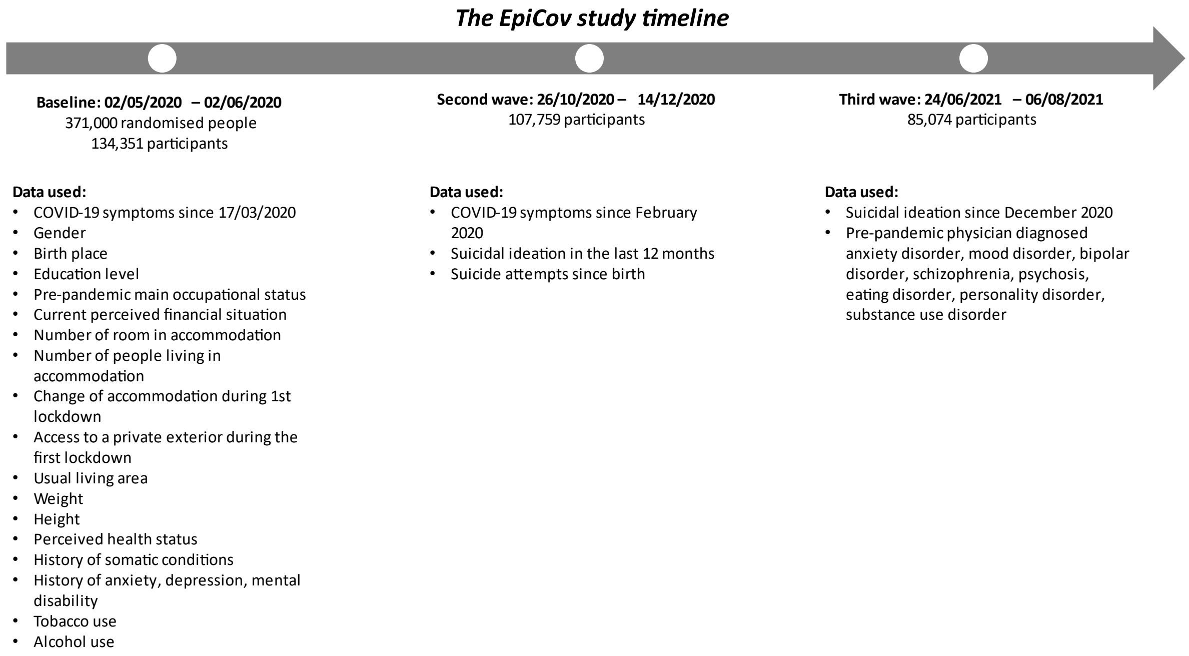

***Tables***

Supplementary Table 1: two-way comparisons of outcome, exposures, and covariates distribution between study sample and excluded sample, weighted by study weights, n = 85,074

| | Included (n = 52,155) | Excluded (n = 32,919) | p |
|---|---|---|---|
| *Suicidal ideations since December 2020* | | | <0.0001 |
| No | 98.32% [98.18 - 98.46] | 89.94% [89.5 - 90.38] | |
| Yes | 1.68% [1.54 - 1.82] | 3.7% [3.44 - 3.97] | |
| Do not wish to answer | .% [. - .] | 6.36% [5.99 - 6.72] | |
| *SARS-CoV2 infection* | | | 0.4217 |
| No | 90.42% [90.08 - 90.75] | 90.94% [89.74 - 92.15] | |
| Yes | 9.58% [9.25 - 9.92] | 9.06% [7.85 - 10.26] | |
| *Covid-19 symptoms between February and December 2020* | | | <0.0001 |
| No | 86.77% [86.39 - 87.14] | 88.38% [87.89 - 88.86] | |
| Yes | 13.23% [12.86 - 13.61] | 11.62% [11.14 - 12.11] | |
| *Gender* | | | <0.0001 |
| Men | 46.13% [45.56 - 46.7] | 49.7% [48.93 - 50.48] | |
| Women | 53.87% [53.3 - 54.44] | 50.3% [49.52 - 51.07] | |
| *Age (years)* | | | <0.0001 |
| 15 - 25 | 11.25% [10.89 - 11.61] | 17.8% [17.23 - 18.37] | |
| 26 - 45 | 27.8% [27.3 - 28.29] | 31.09% [30.38 - 31.79] | |
| 46 - 65 | 34.93% [34.41 - 35.45] | 29.22% [28.56 - 29.88] | |
| > 65 | 26.02% [25.46 - 26.58] | 21.89% [21.16 - 22.63] | |
| *Place of birth* | | | <0.0001 |
| Missing | 1.47% [1.3 - 1.63] | 2.41% [2.13 - 2.68] | |
| Both participant and parents born in mainland France | 80.7% [80.2 - 81.19] | 69.29% [68.53 - 70.05] | |
| Participant or parents born in oversea territories | 1.93% [1.78 - 2.08] | 4.74% [4.45 - 5.04] | |
| Participant born in France of parents born abroad | 8.49% [8.16 - 8.83] | 10.28% [9.81 - 10.75] | |
| Participant born abroad | 7.41% [7.05 - 7.78] | 13.27% [12.63 - 13.92] | |
| *Highest educational attainment* | | | <0.0001 |
| None | 8.23% [7.78 - 8.68] | 15.13% [14.42 - 15.84] | |
| Lower secondary school certificate | 13.3% [12.83 - 13.77] | 16.11% [15.46 - 16.75] | |
| Professional certificate | 19.16% [18.71 - 19.62] | 21.63% [21.01 - 22.25] | |
| Baccalauréat (higher secondary school certificate) | 19.54% [19.11 - 19.98] | 20.22% [19.64 - 20.79] | |
| Bachelor degree or equivalent | 25.31% [24.87 - 25.75] | 18.37% [17.87 - 18.86] | |
| Master degree or more | 14.46% [14.12 - 14.79] | 8.55% [8.23 - 8.86] | |

*weighted % [95% Confidence interval]);*
*chi-square test*

Supplementary Table 1 continued:

| | Included (n = 52,155) | Excluded (n = 32,919) | p |
|---|---|---|---|
| Occupational grade | | | <0.0001 |
| Missing | .% [. - .] | 0.04% [0 - 0.08] | |
| Employed | 47.99% [47.42 - 48.56] | 43.84% [43.09 - 44.6] | |
| Students | 8.76% [8.45 - 9.06] | 12.93% [12.46 - 13.39] | |
| Unemployed | 4.62% [4.35 - 4.89] | 7.51% [7.07 - 7.94] | |
| Retired | 30.36% [29.79 - 30.92] | 23.46% [22.74 - 24.17] | |
| Other including housemakers | 8.27% [7.92 - 8.62] | 12.22% [11.65 - 12.8] | |
| Do not wish to answer | 0% [0 - 0.01] | 0.01% [0 - 0.02] | |
| *Perceived financial situation* | | | <0.0001 |
| Missing | 0.17% [0.1 - 0.24] | 0.39% [0.26 - 0.53] | |
| Comfortable | 16.06% [15.7 - 16.43] | 10.36% [9.95 - 10.77] | |
| Decent | 43.19% [42.63 - 43.75] | 35.86% [35.14 - 36.58] | |
| Short | 30.94% [30.38 - 31.49] | 36.04% [35.29 - 36.8] | |
| Difficult or unbearable without taking loans | 9.44% [9.04 - 9.84] | 16.95% [16.31 - 17.59] | |
| Do not wish to answer | 0.2% [0.12 - 0.27] | 0.4% [0.26 - 0.54] | |
| *Household structure* | | | <0.0001 |
| Missing | 0.07% [0.03 - 0.11] | 0.18% [0.09 - 0.27] | |
| Single | 16.72% [16.22 - 17.21] | 18.35% [17.67 - 19.03] | |
| Couple without children | 32.12% [31.59 - 32.65] | 23.97% [23.33 - 24.6] | |
| Couple with children | 28.7% [28.22 - 29.19] | 25.48% [24.84 - 26.12] | |
| Single-parent | 7.21% [6.9 - 7.52] | 10.62% [10.13 - 11.11] | |
| Participant living with parents | 7.86% [7.55 - 8.16] | 11.83% [11.36 - 12.3] | |
| Complex household | 7.32% [7.01 - 7.63] | 9.58% [9.1 - 10.05] | |
| *Household income per consumption units* | | | <0.0001 |
| Missing | 4.35% [4.05 - 4.64] | 5.14% [4.76 - 5.53] | |
| Less resourceful | 13.13% [12.68 - 13.57] | 22.89% [22.18 - 23.6] | |
| Medium-low | 16.03% [15.53 - 16.53] | 20.73% [20.04 - 21.43] | |
| Medium | 19.1% [18.64 - 19.55] | 19.62% [19 - 20.24] | |
| Medium-high | 22.7% [22.26 - 23.15] | 17.15% [16.65 - 17.66] | |
| Wealthiest | 24.7% [24.29 - 25.11] | 14.46% [14.06 - 14.85] | |

*weighted % [95% Confidence interval];*
*chi-square test*

Supplementary Table 1 continued:

| | Included (n = 52,155) | Excluded (n = 32,919) | p |
|---|---|---|---|
| *Less than one room per person in usual accommodation* | | | <0.0001 |
| Missing | 0.36% [0.28 - 0.43] | 0.59% [0.44 - 0.73] | |
| No | 93.11% [92.79 - 93.43] | 87.26% [86.69 - 87.83] | |
| Yes | 6.54% [6.22 - 6.85] | 12.16% [11.6 - 12.71] | |
| *Residence not in usual housing during the first lockdown* | | | 0.4271 |
| No | 95.03% [94.78 - 95.27] | 95.23% [94.92 - 95.54] | |
| Yes | 4.97% [4.73 - 5.21] | 4.77% [4.46 - 5.08] | |
| Do not wish to answer | 0% [0 - 0.01] | 0% [0 - 0.01] | |
| *Access to a private exterior during the first lockdown* | | | <0.0001 |
| Missing | .% [. - .] | 0% [0 - 0.01] | |
| No | 8.66% [8.31 - 9.02] | 12.83% [12.25 - 13.41] | |
| Yes | 90.38% [90.01 - 90.75] | 85.5% [84.89 - 86.11] | |
| Other situations | 0.93% [0.8 - 1.06] | 1.6% [1.36 - 1.84] | |
| Do not wish to answer | 0.03% [0.01 - 0.04] | 0.06% [0.01 - 0.12] | |
| *Usual area of residence* | | | <0.0001 |
| Less affected area | 65.68% [65.14 - 66.22] | 63.6% [62.85 - 64.36] | |
| Grand-Est | 8.69% [8.39 - 8.99] | 8.02% [7.6 - 8.45] | |
| Hauts-de-France | 8.35% [8.03 - 8.68] | 9.31% [8.86 - 9.76] | |
| Ile-de-France | 17.28% [16.85 - 17.71] | 19.06% [18.43 - 19.69] | |
| *Urban density of living area (urban units)* | | | <0.0001 |
| Oversea territories | 1.26% [1.15 - 1.37] | 3.58% [3.35 - 3.82] | |
| Less than 2,000 | 24.04% [23.56 - 24.52] | 20.79% [20.18 - 21.4] | |
| Between 2,000 and 1,999,999 | 59.61% [59.05 - 60.17] | 58.85% [58.09 - 59.61] | |
| Paris area | 15.09% [14.68 - 15.5] | 16.78% [16.17 - 17.38] | |
| *Usual residence in deprived neighborhood* | | | <0.0001 |
| No | 95.69% [95.39 - 95.98] | 90.81% [90.26 - 91.36] | |
| Yes | 4.31% [4.02 - 4.61] | 9.19% [8.64 - 9.74] | |

*weighted % [95% Confidence interval]; chi-square test*

Supplementary Table 1 continued:

| | Included (n = 52,155) | Excluded (n = 32,919) | p |
|---|---|---|---|
| Perceived general health status | | | <0.0001 |
| Missing | 0.11% (0.06; 0.17) | 0.14% (0.07; 0.21) | |
| Good to very good | 77.7% (77.16; 78.23) | 75.22% (74.49; 75.94) | |
| Quite good | 18.67% (18.17; 19.17) | 19.61% (18.95; 20.26) | |
| Poor to very poor | 3.52% (3.26; 3.78) | 5.02% (4.6; 5.44) | |
| Do not wish to answer | .% (.; .) | 0.01% (0; 0.03) | |
| *Body Mass Index (kg/m²)* | | | <0.0001 |
| Missing | 0.99% (0.84; 1.13) | 2.41% (2.11; 2.71) | |
| Under 18.5 | 3.31% (3.12; 3.51) | 4.51% (4.2; 4.82) | |
| From 18.5 to under 25 | 50.48% (49.91; 51.06) | 48.12% (47.34; 48.89) | |
| From 25 to under 30 | 30.49% (29.96; 31.02) | 29.59% (28.88; 30.3) | |
| From 30 and over | 14.72% (14.29; 15.16) | 15.37% (14.8; 15.94) | |
| *Pre pandemic somatic chronic condition* | | | <0.0001 |
| Missing | 0.07% (0.02; 0.11) | 0.1% (0.04; 0.17) | |
| No | 60.88% (60.3; 61.45) | 64.9% (64.14; 65.66) | |
| Yes | 39.05% (38.48; 39.63) | 34.99% (34.23; 35.75) | |
| Do not wish to answer | 0% (0; 0.01) | 0.01% (0; 0.02) | |
| Pre pandemic mental health disorder | | | <0.0001 |
| Missing | 0.05% (0.01; 0.09) | 0.1% (0.03; 0.16) | |
| No | 88.94% (88.56; 89.33) | 86.91% (86.38; 87.43) | |
| Yes | 11% (10.62; 11.38) | 12.99% (12.47; 13.51) | |
| Do not wish to answer | 0% (0; 0.01) | 0.01% (0; 0.01) | |
| *Tobacco use* | | | <0.0001 |
| Missing | 0.27% (0.19; 0.36) | 0.57% (0.42; 0.72) | |
| Never | 48.66% (48.08; 49.23) | 51.77% (51; 52.54) | |
| Past only | 32.15% (31.63; 32.68) | 23.94% (23.32; 24.57) | |
| Current | 18.86% (18.39; 19.32) | 23.66% (22.99; 24.33) | |
| Do not wish to answer | 0.06% (0.03; 0.09) | 0.05% (0.03; 0.08) | |
| *Alcohol use* | | | <0.0001 |
| Missing | 0.27% (0.19; 0.35) | 0.58% (0.43; 0.73) | |
| Never | 29.3% (28.75; 29.86) | 41.63% (40.84; 42.41) | |
| Rare | 13.56% (13.16; 13.96) | 14.69% (14.14; 15.25) | |
| Occasional | 23.3% (22.84; 23.77) | 19.47% (18.9; 20.05) | |
| Often | 23.04% (22.59; 23.48) | 15.62% (15.13; 16.11) | |
| Daily | 10.52% (10.17; 10.87) | 7.97% (7.57; 8.37) | |
| Do not wish to answer | 0% (0; 0.01) | 0.04% (0.01; 0.07) | |
| *Hospitalization rates in place of residence during the 1st lockdown* | | | 0.0006 |
| Missing | 4.86% (4.62; 5.09) | 4.57% (4.28; 4.87) | |
| Lowest | 23.25% (22.76; 23.73) | 22.99% (22.35; 23.63) | |
| Medium-low | 24.75% (24.25; 25.25) | 23.34% (22.7; 23.99) | |
| Medium-High | 22.47% (21.98; 22.95) | 23.15% (22.5; 23.8) | |
| Highest | 24.68% (24.19; 25.17) | 25.94% (25.24; 26.64) | |

*weighted % [95% Confidence interval];*
*chi-square test*

Supplementary table 2: two-way comparisons of exposures, and covariates distribution according to the outcome, n = 52,050

| | Suicidal Ideation | | |
|---|---|---|---|
| | No (51,187) | Yes (863) | p |
| *SARS-CoV2 infection* | | | 0.8610 |
| No | 90.42% [90.09 - 90.76] | 90.65% [88.21 - 93.08] | |
| Yes | 9.58% [9.24 - 9.91] | 9.35% [6.92 - 11.79] | |
| *Covid-19 symptoms between February and December 2020* | | | <0.0001 |
| No | 86.93% [86.55 - 87.31] | 77.39% [74 - 80.78] | |
| Yes | 13.07% [12.69 - 13.45] | 22.61% [19.22 - 26] | |
| *Gender* | | | 0.8326 |
| Men | 46.17% [45.59 - 46.75] | 45.71% [41.48 - 49.94] | |
| Women | 53.83% [53.25 - 54.41] | 54.29% [50.06 - 58.52] | |
| *Age (years)* | | | <0.0001 |
| 15 - 25 | 11.03% [10.67 - 11.38] | 24.42% [20.68 - 28.16] | |
| 26 - 45 | 27.72% [27.22 - 28.23] | 34.1% [30.14 - 38.07] | |
| 46 - 65 | 35.02% [34.49 - 35.54] | 29.09% [25.57 - 32.6] | |
| > 65 | 26.23% [25.66 - 26.8] | 12.39% [9.22 - 15.56] | |
| *Place of birth* | | | 0.4372 |
| Both participant and parents born in mainland France | 81.99% [81.5 - 82.48] | 82.99% [79.74 - 86.25] | |
| Participant or parents born in oversea territories | 1.96% [1.81 - 2.11] | 2.16% [0.56 - 3.77] | |
| Participant born in France of parents born abroad | 8.59% [8.25 - 8.93] | 9.38% [7.1 - 11.66] | |
| Participant born abroad | 7.46% [7.09 - 7.83] | 5.47% [3.38 - 7.56] | |
| *Highest educational attainment* | | | <0.0001 |
| None | 8.26% [7.81 - 8.71] | 4.69% [2.54 - 6.84] | |
| Lower secondary school certificate | 13.32% [12.84 - 13.8] | 12.35% [9.36 - 15.35] | |
| Professional certificate | 19.21% [18.75 - 19.66] | 13.61% [10.64 - 16.57] | |
| Baccalauréat (higher secondary school certificate) | 19.49% [19.05 - 19.92] | 23.24% [19.54 - 26.93] | |
| Bachelor degree or equivalent | 25.3% [24.86 - 25.75] | 27.4% [23.93 - 30.87] | |
| Master degree or more | 14.42% [14.09 - 14.75] | 18.71% [15.64 - 21.78] | |

*weighted % [95% Confidence interval];*
*chi-square test*

Supplementary table 2 continued:

| | Suicidal Ideation | | |
|---|---|---|---|
| | No (51,187) | Yes (863) | p |
| Occupational grade | | | <0.0001 |
| Employed | 47.99% [47.41 - 48.56] | 50.21% [46.02 - 54.4] | |
| Students | 8.54% [8.23 - 8.84] | 21.51% [17.92 - 25.1] | |
| Unemployed | 4.63% [4.36 - 4.91] | 4.46% [2.63 - 6.29] | |
| Retired | 30.62% [30.05 - 31.19] | 15.14% [11.85 - 18.44] | |
| Other including housemakers | 8.23% [7.88 - 8.58] | 8.68% [6.51 - 10.84] | |
| *Perceived financial situation* | | | <0.0001 |
| Comfortable | 16.16% [15.8 - 16.53] | 15.73% [12.73 - 18.73] | |
| Decent | 43.51% [42.94 - 44.08] | 36.52% [32.57 - 40.48] | |
| Short | 30.98% [30.42 - 31.55] | 32.05% [28.14 - 35.97] | |
| Difficult or unbearable without taking loans | 9.35% [8.94 - 9.75] | 15.69% [12.35 - 19.03] | |
| *Household structure* | | | <0.0001 |
| Single | 16.74% [16.24 - 17.24] | 15.73% [12.82 - 18.64] | |
| Couple without children | 32.3% [31.77 - 32.84] | 21.27% [17.7 - 24.85] | |
| Couple with children | 28.81% [28.32 - 29.3] | 25.47% [22.08 - 28.86] | |
| Single-parent | 7.16% [6.85 - 7.47] | 9.88% [7.39 - 12.38] | |
| Participant living with parents | 7.77% [7.46 - 8.07] | 12.62% [9.98 - 15.26] | |
| Complex household | 7.21% [6.91 - 7.52] | 15.02% [11.57 - 18.47] | |
| *Household income per consumption units* | | | 0.0449 |
| Less resourceful | 13.8% [13.33 - 14.26] | 17.99% [14.76 - 21.22] | |
| Medium-low | 16.57% [16.05 - 17.08] | 18.04% [14.51 - 21.57] | |
| Medium | 19.86% [19.38 - 20.33] | 18.3% [14.84 - 21.76] | |
| Medium-high | 23.73% [23.27 - 24.19] | 21.99% [18.6 - 25.38] | |
| Wealthiest | 26.05% [25.62 - 26.48] | 23.68% [20.49 - 26.87] | |

*weighted % [95% Confidence interval];*
*chi-square test*

Supplementary table 2 continued:

| | Suicidal Ideation | | |
|---|---|---|---|
| | No (51,187) | Yes (863) | p |
| *Less than one room per person in usual accommodation* | | | 0.0206 |
| No | 93.39% [93.07 - 93.71] | 90.64% [87.96 - 93.33] | |
| Yes | 6.61% [6.29 - 6.93] | 9.36% [6.67 - 12.04] | |
| *Residence not in usual housing during the first lockdown* | | | <0.0001 |
| No | 95.11% [94.87 - 95.36] | 89.81% [87.22 - 92.4] | |
| Yes | 4.89% [4.64 - 5.13] | 10.19% [7.6 - 12.78] | |
| *Access to a private exterior during the first lockdown* | | | 0.0450 |
| No | 8.62% [8.26 - 8.98] | 11.42% [8.89 - 13.95] | |
| Yes | 90.45% [90.07 - 90.83] | 87.75% [85.15 - 90.35] | |
| Other situations | 0.93% [0.8 - 1.06] | 0.83% [0.15 - 1.51] | |
| Usual living area | | | 0.0026 |
| Less affected area | 65.72% [65.18 - 66.27] | 62.3% [58.2 - 66.39] | |
| Grand-Est | 8.75% [8.44 - 9.06] | 5.96% [4.12 - 7.79] | |
| Hauts-de-France | 8.31% [7.98 - 8.64] | 10.04% [7.53 - 12.55] | |
| Ile-de-France | 17.21% [16.78 - 17.65] | 21.71% [18.11 - 25.3] | |
| *Urban density of living area (urban units)* | | | 0.0172 |
| Oversea territories | 1.27% [1.16 - 1.38] | 1.07% [0.47 - 1.67] | |
| Less than 2,000 | 24.09% [23.6 - 24.58] | 22.74% [19.1 - 26.38] | |
| Between 2,000 and 1,999,999 | 59.63% [59.06 - 60.19] | 56.58% [52.38 - 60.78] | |
| Paris area | 15.02% [14.6 - 15.43] | 19.61% [16.13 - 23.09] | |
| *Usual residence in deprived neighborhood* | | | 0.9169 |
| No | 95.69% [95.39 - 95.99] | 95.78% [94.06 - 97.51] | |
| Yes | 4.31% [4.01 - 4.61] | 4.22% [2.49 - 5.94] | |

*weighted % [95% Confidence interval];*
*chi-square test*

Supplementary table 2 continued:

| | Suicidal Ideation | | |
|---|---|---|---|
| | No (51,187) | Yes (863) | p |
| Perceived general health status | | | 0.0148 |
| Good to very good | 77.89% [77.35 - 78.43] | 72.81% [68.87 - 76.74] | |
| Quite good | 18.65% [18.14 - 19.15] | 22.04% [18.33 - 25.76] | |
| Poor to very poor | 3.46% [3.2 - 3.72] | 5.15% [3.21 - 7.09] | |
| *Body Mass Index (kh/m²)* | | | <0.0001 |
| Under 18.5 | 3.32% [3.12 - 3.52] | 6.75% [4.54 - 8.96] | |
| From 18.5 to under 25 | 50.79% [50.21 - 51.37] | 51.67% [47.47 - 55.87] | |
| From 25 to under 30 | 30.98% [30.44 - 31.52] | 25.68% [22.02 - 29.35] | |
| From 30 and over | 14.91% [14.47 - 15.35] | 15.89% [12.65 - 19.14] | |
| *Pre pandemic somatic condition* | | | 0.7645 |
| None | 60.92% [60.34 - 61.5] | 60.3% [56.25 - 64.34] | |
| At least one | 39.08% [38.5 - 39.66] | 39.7% [35.66 - 43.75] | |
| Pre pandemic mental health disorder | | | <0.0001 |
| None | 89.4% [89.02 - 89.79] | 65.27% [61.31 - 69.23] | |
| At least one | 10.6% [10.21 - 10.98] | 34.73% [30.77 - 38.69] | |
| *Tobacco use* | | | 0.0001 |
| Never | 48.91% [48.33 - 49.49] | 41.36% [37.24 - 45.47] | |
| Past only | 32.25% [31.72 - 32.78] | 33.46% [29.5 - 37.41] | |
| Current | 18.83% [18.36 - 19.3] | 25.19% [21.51 - 28.87] | |
| *Alcohol use* | | | 0.4830 |
| Never | 29.39% [28.82 - 29.95] | 29.08% [25.2 - 32.97] | |
| Rare | 13.57% [13.17 - 13.98] | 14.45% [11.36 - 17.54] | |
| Occasional | 23.39% [22.92 - 23.86] | 20.44% [17.13 - 23.74] | |
| Often | 23.11% [22.66 - 23.56] | 24.2% [20.7 - 27.7] | |
| Daily | 10.54% [10.18 - 10.89] | 11.83% [9.22 - 14.44] | |
| *Hospitalization rates in place of residence during the 1st lockdown* | | | 0.7270 |
| Lowest | 23.63% [23.13 - 24.12] | 23.46% [19.89 - 27.03] | |
| Medium-low | 27.22% [26.7 - 27.74] | 25.25% [21.82 - 28.68] | |
| Medium-High | 23.41% [22.92 - 23.9] | 24.15% [20.42 - 27.88] | |
| Highest | 25.74% [25.24 - 26.24] | 27.14% [23.37 - 30.91] | |

*weighted % [95% Confidence interval];*
*chi-square test*

***Directed Acyclic Graph (DAG)***

A DAG helps visualizing the different pathways through which the exposure, outcome and covariates are related to each other. It also gives the minimum adjustment required to assess the direct relationship between the exposure and outcome, based on the specified pathways and covariates. For the present study, we only used a DAG for visualization as decided to base our covariate selection on current available literature.

*Supplementary figure 2: directed acyclic graph of the links between COVID-19 disease, suicidal ideation and all identified covariates*

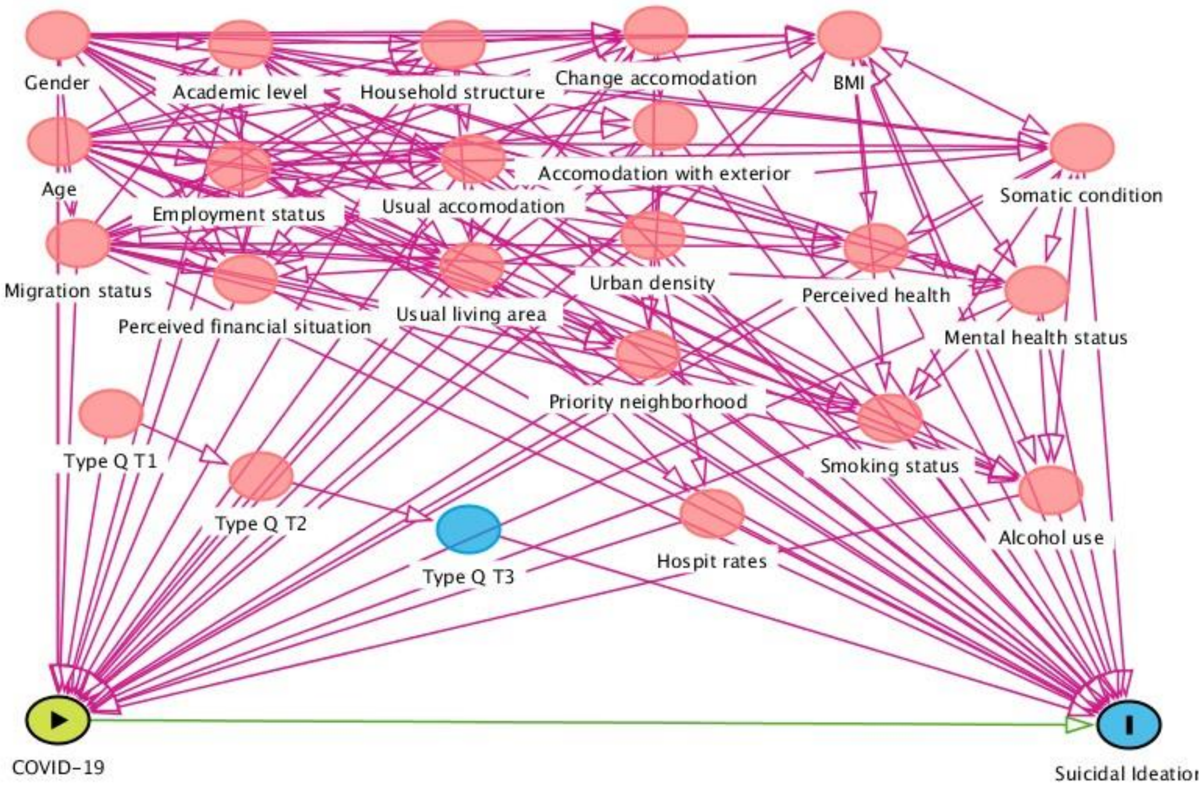


***Multiple imputation***

We used raw covariates as much as possible, with “do not wish to answer” and “do not know” modalities when applicable. 24 covariates were used in the multiple imputation model, with 13 of them having up to 4.71% missing data. The seed was randomly define as 91118. Below is a screen capture of the SAS code used, followed by the matching covariate definitions:

*Supplementary figure 3: screen capture of the SAS code used for imputation analyses*

```
proc mi data=SuicidCovid_IM_All out=CDPbase.SuicidCovid_IM_All nimpute=5 seed=91118;
    class t1_typq sexe dipln t1_a7n t1_a12n t1_c2n denspop_cdp qpv_insee t1_b7n t1_b6n typfam t1_c19n t1_b5n t1_i4n t1_i2n
        ori5cl nviedec lieunais t1_deptconf;
    fcs logistic (t1_b6n typfam t1_c19n t1_b5n t1_i4n t1_i2n
                    ori5cl nviedec lieunais / likelihood=augment)
        discrim (t1_b7n t1_deptconf / CLASSEFFECTS=include)
        reg (t1_logdens t1_poids t1_taille);
    var t1_typq age_insee sexe dipln t1_a7n t1_a12n t1_c2n denspop_cdp qpv_insee t1_b7n t1_b6n typfam t1_c19n t1_b5n t1_i4n t1_i2n t1_logdens
        t1_poids t1_taille ori5cl nviedec lieunais t1_deptconf;
run;
```

*Supplementary table 3: definition of all covariates used for imputation*

| | Definition |
|---|---|
| t1_typq | Questionnaire type at first data collection time point (CATI/CAWI) |
| age_insee | Age according to INSEE |
| sexe | You are… (a man/a woman + do not wish to answer) |
| dipln | What degree(s) do you have? (List of 13 possibilities) |
| t1_a7n | During lockdown did you live most of the time in your usual accommodation (main residence)? (yes/no + do not wish to answer) |
| t1_a12n | Is this lockdown accommodation: (in an apartment without balcony, terrace or collective garden/in an apartment with a balcony or terrace/in an apartment with a collective garden/in a house with no yard or garden/in a house with a yard or garden/in a dwelling of fortune/other + do not wish to answer) |
| t1_c2n | What was your main occupation before the lockdown began? (Employment (salaried or self-employed, including helping someone with their job)/Apprenticeship under contract or paid internship/Studies (pupil, student) or unpaid internship/Unemployment (registered or not at the Pôle Emploi)/Retirement or early retirement (former employee or former self-employed)/Housewife or man/Other situation + do not wish to answer) |
| denspop_cdp | Urban density based on data from INSEE |
| qpv_insee | Usual accommodation in a deprived neighborhood according to INSEE |
| t1_ind_poids | Index of answer to weight (had answered/do not know/do not wish to answer) |
| t1_ind_taille | Index of answer to height (had answered/do not know/do not wish to answer) |
| t1_b7n | Have you been limited, for at least 6 months, because of a health problem, in the activities people usually do? (yes, strongly limited/yes, limited but not strongly/no/do not wish to answer) |
| t1_b6n | Do you have an illness or health problem that is chronic or long-lasting? (yes/no/do not wish to answer) |
| typfam | Household structure according to INSEE |
| t1_c19n | Financially in your household would you rather say that today… (You are comfortable/It's okay/It's tight, you have to be careful/You can hardly do it/You can't do it without going into debt (or using consumer credit) + do not wish to answer) |
| t1_b5n | How is your general health? (Very good/Good/Pretty good/Bad/Very bad + do not wish to answer) |
| t1_i4n | Do you currently drink alcoholic beverages i.e. wine, beer, hard liquor or other alcohols such as cider, port, champagne? (every day/four to six times of week/two to three times a week/once a week/once a week or more but no further detail/once a month or more/less than that/never + do not wish to answer) |
| t1_i2n | Do you currently smoke tobacco (cigarettes, cigars, cigarillos, pipe, hookah or shisha), even if only from time to time (apart from electronic cigarettes)? (currently smoking/past daily smoker for six months or more/past daily smoker for less than 6 months/pas occasional smoker/smoked only once or twice to try/never smoked + do not wish to answer) |
| t1_logdens | number of room in accommodation during lockdown (excluding bathroom and kitchen) / number of people living in this accommodation |
| t1_poids | weight in kilograms |
| t1_taille | Height in centimeters |
| ori5cl | Origins based on participants answer (majority population/born in oversea territories or from parents born in oversea territories/ born in mainland France from parents born outside of France/born outside of France) |
| lieunais | Origins according to INSEE(French/French from oversea department/French from oversea communes/Unavailable/Strangers) |

t1_deptconf French department of accommodation during the first lockdown

***Inverse Probability weighting (IPW) method***

In the present work, IPW was used as an adjustment strategy to assess the direct relationship between COVID-19 infection in 2020 and subsequent suicidal ideation in the first half of 2021. Two binary COVID-19 infection markers were separately used, self-reported COVID-19 symptoms (yes or no) and SARS-CoV2 serology (positive or negative). Participants reporting COVID-19 symptoms or being seropositive will be referred to as cases below. First, propensity scores, i.e. probabilities (p), of either self-reporting COVID-19 symptoms or being seropositive to SARS-CoV2 were computed using a logistic regression where the COVID-19 markers were explained by the covariates listed in the manuscript. Then inverse probability weights were calculated as follow:

- In cases: ipw = $\frac{1}{(1-p)}$
- In control participants: ipw = $\frac{1}{p}$

Where p is the probability of self-reporting symptoms, or being seropositive, according to the covariates.

IPW is an average treatment effect methodology where the weights are used to create a subpopulation were both the case and control groups have the same covariate distributions as the whole population.

***Average treatment effect on the treated method***

To test robustness of our results we assessed what would have happened to participants classified as cases if they had been classified as controls. To do so, we weighted the control group to have the same covariate distributions as the case group. Conservation of results as compare to IPW method is in favor of true “impact” of the exposition. As for ipw, propensity scores were used to calculate relevant weights:

- In cases: watt = 1
- In control participants: watt = $\frac{p}{(1-p)}$

Where p is the probability of self-reporting symptoms, or being seropositive, according to the covariates.

***Covariates used for propensity scores calculation***

All covariates listed in the manuscript were used to compute the scores. Age was squared and used as a continuous covariate.

*Supplementary Figure 4 : COVID-19 illness distribution, Cohen's distance, and variance ratio before and after IPWeighting, n = 52,050*

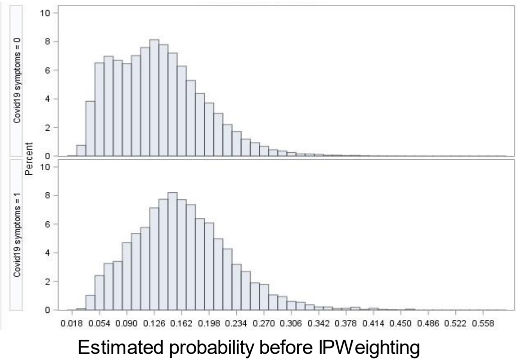


| | \|d\| (%) | Variance ratio |
|---|---|---|
| Before IPW | 52·86 | 0·82 |
| After IPW | 9·95 | 0·95 |

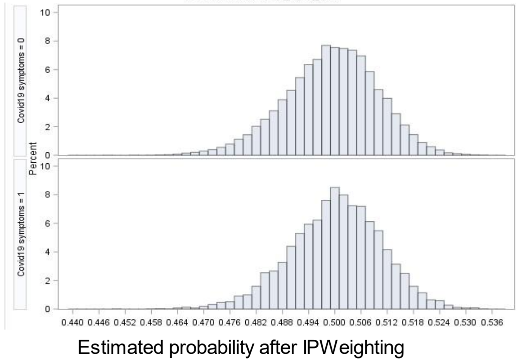


*Supplementary Figure 5 : SARS-CoV2 infection distribution, Cohen's distance, and variance ratio before and after IPWeighting, n = 52,050*

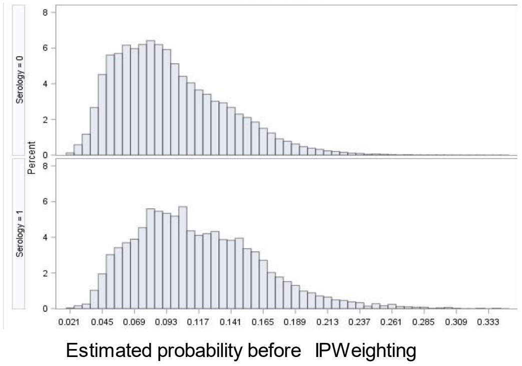


| | \|d\| (%) | Variance ratio |
|---|---|---|
| Before IPW | 46·95 | 0·98 |
| After IPW | 2·72 | 1·06 |

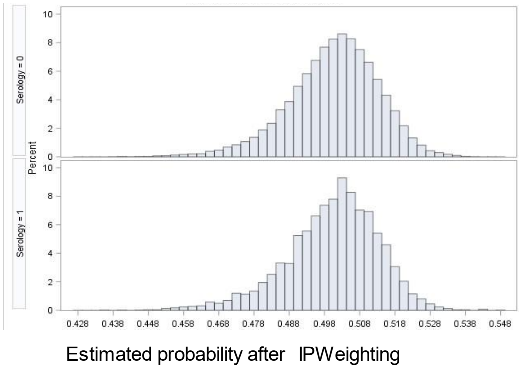

Supplementary table 4: standardized mean differences and p-value of chi-square or student t-test between covariates and COVID-19 illness, before and after IPWeighting

| | COVID-19 illness | | | |
|---|---|---|---|---|
| | Before IPWeighting | | After IPWeighting | |
| | \|SMD\| (%) | p | \|SMD\| (%) | p |
| *Gender* | 10.53 | *** | 0.30 | ns |
| *Place of birth* | 3.95 | *** | 0.92 | ns |
| *Highest educational attainment* | 6.83 | *** | 0.86 | * |
| *Occupational grade* | 14.76 | *** | 0.40 | ns |
| *Perceived financial situation* | 6.75 | *** | 0.28 | ns |
| *Household structure* | 9.28 | *** | 0.18 | ns |
| *Household income per consumption units* | 3.28 | *** | 0.32 | ns |
| *Residence not in usual housing during the first lockdown* | 7.23 | *** | 0.36 | ns |
| *Less than one room per person in usual accommodation* | 11.92 | *** | 0.06 | ns |
| *Access to a private exterior during the first lockdown* | 4.57 | *** | 0.36 | ns |
| *Usual living area* | 11.00 | *** | 0.28 | ns |
| *Urban density of living area (urban units)* | 6.91 | *** | 0.62 | ns |
| *Usual residence in deprived neighborhood* | 3.41 | ** | 0.68 | ns |
| *Perceived general health status* | 6.49 | *** | 0.13 | ns |
| *Body Mass Index (kg/m²)* | 3.11 | *** | 0.21 | ns |
| *Pre pandemic somatic condition* | 0.78 | ns | 0.83 | ns |
| *Pre pandemic mental health disorder* | 13.64 | *** | 0.59 | ns |
| *Tobacco use* | 4.73 | *** | 0.41 | ns |
| *Alcohol use* | 2.76 | *** | 0.54 | ns |
| *Hospitalization rates in place of residence during the 1st lockdown* | 10.23 | *** | 0.57 | ns |
| *Age (squared)* | 37.97 | *** | 0.20 | ns |

*|SMD| (%): absolute value of standardized mean difference in percentage*

*p: p-value of chi-square test or student t-test between each covariate and the relevant exposure*

**: 0.01<p-value≤0.05*

***: 0.001<p-value≤0.01*

****: p-value≤0.001*

Supplementary table 5: standardized mean differences and p-value of chi-square or student t-test between covariates and SARS-CoV2 infection, before and after IPWeighting

| | SARS-CoV2 infection | | | |
|---|---|---|---|---|
| | Before IPWeighting | | After IPWeighting | |
| | \|SMD\| (%) | p | \|SMD\| (%) | p |
| *Gender* | 3.66 | * | 0.41 | ns |
| *Place of birth* | 8.75 | *** | 0.89 | ns |
| *Highest educational attainment* | 4.76 | *** | 0.87 | * |
| *Occupational grade* | 9.93 | *** | 0.59 | ns |
| *Perceived financial situation* | 2.22 | * | 0.97 | * |
| *Household structure* | 9.08 | *** | 0.64 | ns |
| *Household income per consumption units* | 1.66 | ns | 0.98 | * |
| *Residence not in usual housing during the first lockdown* | 7.47 | *** | 0.39 | ns |
| *Less than one room per person in usual accommodation* | 11.08 | *** | 0.28 | ns |
| *Access to a private exterior during the first lockdown* | 3.81 | *** | 0.25 | ns |
| *Usual living area* | 13.38 | *** | 0.28 | ns |
| *Urban density of living area (urban units)* | 9.21 | *** | 0.27 | ns |
| *Usual residence in deprived neighborhood* | 7.56 | *** | 1.29 | * |
| *Perceived general health status* | 8.59 | *** | 0.56 | ns |
| *Body Mass Index (kg/m²)* | 2.92 | * | 0.54 | ns |
| *Pre pandemic somatic condition* | 12.29 | *** | 0.16 | ns |
| *Pre pandemic mental health disorder* | 4.98 | ** | 0.71 | ns |
| *Tobacco use* | 12.76 | *** | 0.65 | ns |
| *Alcohol use* | 3.81 | *** | 0.62 | ns |
| *Hospitalization rates in place of residence during the 1st lockdown* | 13.74 | *** | 0.23 | ns |
| *Age (squared)* | 24.62 | *** | 0.11 | ns |

*|SMD| (%): absolute value of standardized mean difference in percentage*

*p: p-value of chi-square test or student t-test between each covariate and the relevant exposure*

**: 0.01<p-value≤0.05*

***: 0.001<p-value≤0.01*

****: p-value≤0.001*
